# Tracking SARS-CoV-2 RNA through the wastewater treatment process

**DOI:** 10.1101/2020.10.14.20212837

**Authors:** Hala Abu Ali, Karin Yaniv, Edo Bar-Zeev, Sanhita Chaudhury, Marilou Shaga, Satish Lakkakula, Zeev Ronen, Ariel Kushmaro, Oded Nir

## Abstract

The municipal sewage carries the new coronavirus (SARS-CoV-2), shed by COVID-19 patients, to wastewater treatment plants. Proper wastewater treatment can provide an important barrier for preventing uncontrolled discharged of the virus into the environment. However, the role of the different wastewater treatment stages in reducing virus concentrations was, thus far, unknown. In this work, we quantified SARS-CoV-RNA in the raw sewage and along the main stages of the wastewater process from two different plants in Israel during this COVID-19 outbreak. We found that ca. 2 Log removal could be attained after primary and secondary treatment. Despite this removal, significant concentrations of SARS-CoV-RNA (>100 copies per mL) could still be detected in the treated wastewater. However, after treatment by chlorination, SARS-CoV-RNA was detected only once, likely due to insufficient chlorine dose. Our results highlight the need to protect wastewater treatment plants operators, as well as populations living near areas of wastewater discharge, from the risk of infection. In addition, our results emphasize the capabilities and limitations of the conventional wastewater treatment process in reducing SARS-CoV-RNA concentration, and present preliminary evidence for the importance of tertiary treatment and chlorination in reducing SARA-CoV-2 dissemination.

## INTRODUCTION

The new human coronavirus (SARS-CoV-2), was found in urine and feces ^1–9^ of infected individuals, as well as in various wastewater systems.^6,10–14^ Consequently, there has been a recent global interest in monitoring the dissemination of SARS-CoV-2 in municipal sewages, mainly for wastewater-based epidemiology (WBE). ^3,15–20^ The implication that wastewater is a potential dissemination and infection route, has been also discussed,^13,21–23^however, empirical evidences are still scarce. This is partially due to the current challenge of sample concentration and extraction methods,^24^ required for precise virus quantification.

Recent studies have shown that SARS-CoV-2 RNA could be found in the outlet of wastewater treatment plants (WWTP), as well as in water bodies receiving the treated WW,^12,16,25^ indicating a public health risk via the faecal-oral or faecal-aerosol infection routes.^2,26–28^ Indeed, a recent study showed 90% removal of SARS-CoV-2 infectivity only after 1.5 days in wastewater.^29^ This is longer than the hydraulic residence time in many urban pipelines^30^ and longer than typical hydraulic residence time (HRT) in WWTPs. Therefore the risk of infection through contact (direct or through aerosols) with conventionally treated wastewater, cannot be dismissed at this point, especially where treated wastewater is reused for agriculture, or recreation near urban areas.^31^ Although the extent of infectivity associated with SARS-CoV-2 RNA in treated WW is not yet clear,^11,31^ this unnecessary risk can be minimized by ensuring an almost complete viral RNA removal by the WWTP.

The fate of SARA-CoV-2in WWTPs, similarly to other viruses,^8^ is expected to be influenced by various operational and environmental factors. Conventional WWTP includes several steps in series (Figure 1), ^8^ each step contributing to the removal of the virus from the water through physical, biological or chemical mechanisms.^31^ The virus removal rate or the neutralization of its infectivity at each step depends on the plant’s operating conditions such as wastewater and biomass residence times,^32^ as well as on environmental factors such as temperature, pH and organic matter.^26,31,32^ Therefore, it is hard to rule out the risk of infection via WW, even though recent preliminary tests suggest that SARS-CoV-2 particles in treated WW are not infective.^11^ Given this uncertainty and the extent of the pandemic, it is critical to determine the fate of the virus at the outlets, as well as in the different treatment stages of WWTP’s. This information may be crucial for regulating and adapting the WW treatment sector to the COVID-19 pandemic, mainly with respect to: (1) assessing the risk of viral dissemination by treated WW. (2) Recognizing the critical treatment stages required for preventing viral dissemination – with implications on maintenance and future development of WWTP’s. (3) Ensuring the health and safety of plant workers, and the general population exposed to the treated WW.

**Figure 1.**
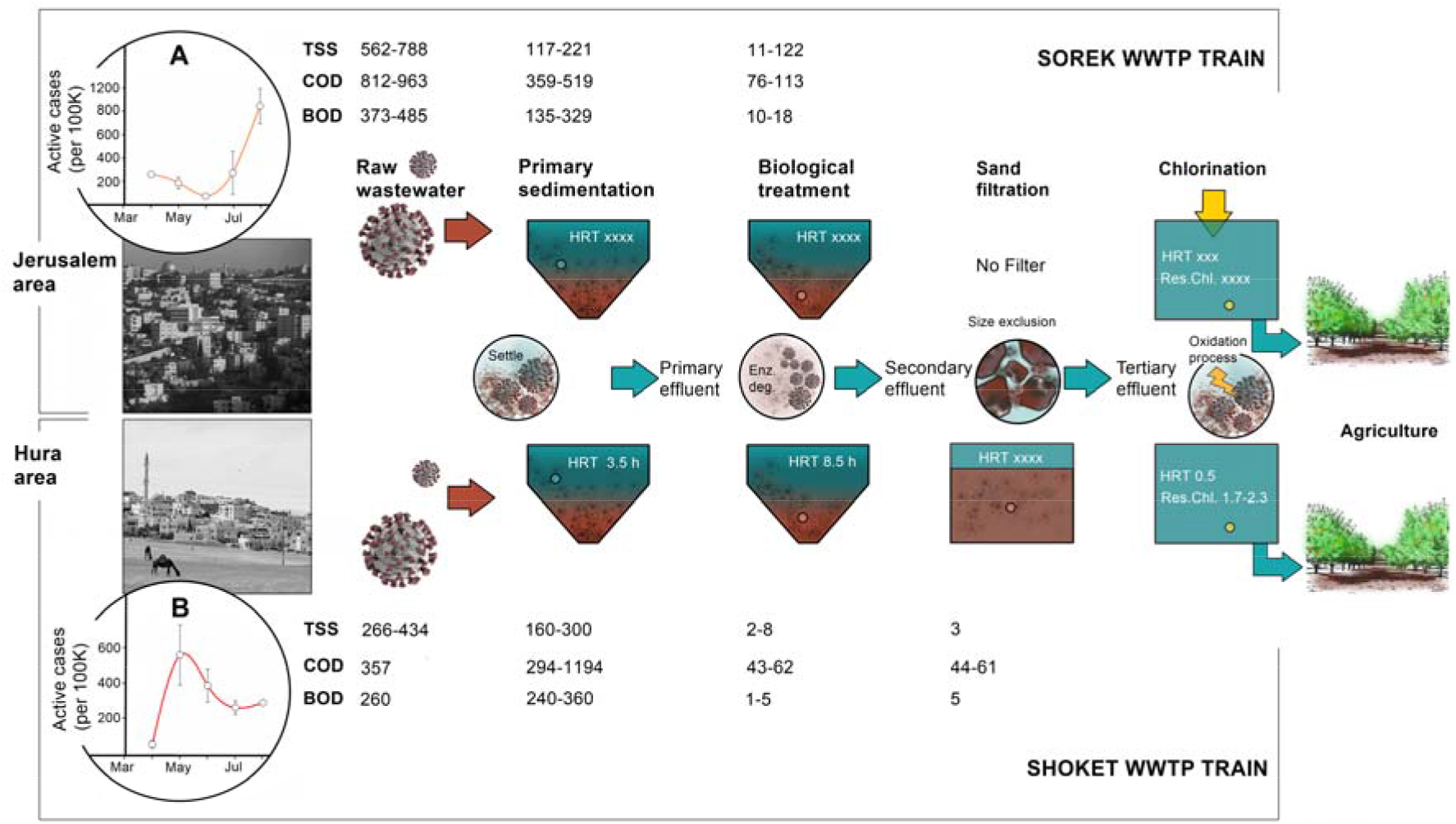
Illustration of the different stages involved in the WW treatment process in A) Sorek and B) Shoket WWTP. The time dependent active cases of covid-19 cases per 100 K residents in the Jerusalem and the Hura region are also shown to highlight the variability between different times (twice in April end and once in July) and locations in the context of WW sampling. The TSS, COD and BOD data (corresponding to different treatment stages) are shown to indicate on the process performances of the WWTPs. In the activated sludge process, the primary effluent is collected after the 1^st^ step, *i*.*e*., the sedimentation step, where the raw WW is settled to remove large solids. This is followed by aerobic biological treatment that reduces organic content and a secondary sedimentation step that separate the secondary (activated) sludge from the secondary effluent. Sand filtration is applied as a tertiary treatment in Shoket WWTP only. Before exiting the WWTP the effluent in both locations is subjected to disinfection through chlorination. The disinfected effluent is collected in irrigation reservoirs and then reused in agriculture.

Nevertheless, to-date there is no information (to the best of our knowledge) on the presence of SARS-CoV-2 along the treatment stages, while data on the virus presence at the outlet is scarce. Therefore, in the present study, we present the first dataset on SARS-CoV-2 (RNA) presence along the different treatment stages, from inlet to outlet, in two different WWTPs in Israel. Our results highlight the importance of proper WWTP operation for obtaining high virus removal rate and stress the need for more research and better risk assessments to protect WWTP workers and prevent dissemination through WW.

## MATERIALS AND METHODS

The details of all the materials used in this work, are given in the supporting information (SI, section A). Most relevant protocols are shortly described herein, while more details are given in the SI when needed. For all the protocols, whenever needed, aappropriate protection gear (masks, hands protection, coveralls) was used while handling the samples in a biosafety level 2 (BSL-2) biological hood.

### 2.1. Sewage Sampling and further processing

Grab samples were collected from two different WWTPs in Israel - Sorek and Shoket, treating WW from the areas of Jerusalem and Hura respectively. The WW samples (volumes in SI, section C1) were collected in a sanitized and sterilized disposable plastic sampling container. Post sample collection, the containers were externally sprayed with 2% sodium hypochlorite and 70% ethanol, inserted into pre-labelled biohazard bags. After sealing, these biohazard bags were sprayed with disinfectants for further sterilization. The WW samples were then transported in an ice cooler to the laboratory. Upon arrival, the samples were re-sterilized using the previous methodology. Immediately after collection, the samples were stored at 4 °C for no longer than 24 h.^33^

Apart from the raw WW, samples (initial volumes in SI, section C1) were concentrated to 200 mL by ultrafiltration using sterile dialysis filters with an average pore size of 3.3 nm and an active surface area of 2.2 m^2^ (Fresenius, Bud Hamburg, Germany)^34,35^ and a peristaltic pump with new cleaned tubing that was replaced every time. A similar method was previously described in Obayomi et al., 2019.^36^ After concentrating, the accumulated particles in the filter were eluted with Phosphate Buffered Saline (PBS). More details can be found in the SI (section A).

### 2.2 RNA extraction and detection/quantification of SARS-CoV

RNA extractions were performed using 200 µL of raw or concentrated WW samples using NucleoSpin RNA (Macherey Nagel, Germany). RNA was eluted with 50 µL of RNase free water and kept at –80°C. For SARS-CoV-2 detection, we used One Step PrimeScript III RT-qPCR mix (TAKARA, Japan) with CDC’s primers and probe N1 together with 5 µL of eluted RNA. RT-qPCR amplification was executed using Step One Plus real-time PCR system (Applied Biosystems, Thermo Scientific). For quality control in each RT-PCR run, positive (N gene plasmid, IDT) and non-template control (NTC) were performed. Copy number calculation was done according to standard calibration curve as detailed in SI (section C2).

## RESULTS AND DISCUSSION

We tracked the presence of SARS-CoV-2 RNA on three different dates and in two separate wastewater treatment plants (Figure1). Samples were collected twice during April 2020, in the midst of the ‘first wave’ of the COVID19 outbreak in Israe;l^37^ and once in July 2020, at the beginning of Israel’s ‘second wave’. Sorek WWTP treats the sewage of west Jerusalem municipality, a densely populated area of the city that experienced similar trend of infection as observed in Israel in March to July (Figure 1A). Shoket WWTP, (Figure 1B) — the 2^nd^ sampling site mainly treats sewage originating from *Hebron* city in the *West Bank*. The sewage is discharged in the Hebron ephemeral stream and makes up ca. two thirds of the inflow to Shoket WWTP. In addition, Shoket treats the sewage of a rural area in the southern part of Israel that includes Hura, Meitar and other small municipalities. Specifically, Hura suffered from high infection rates during the 1^st^ COVID-19 wave in Israel and a lower one in the second wave, whereas the West Bank suffered from a surge of confirmed COVID-19 cases during July.^38^ Both WWTPs operate a conventional activated sludge process ^39–41^(Figure 1), with a primary settler followed by a secondary treatment that includes an aerobic biological reactor and a secondary settler to separate the biomass. Overall, the data (Figure 1 and Table S1) provided by the plant’s operator indicate a proper and stable performance of the secondary treatment with COD/BOD average removal rates (%) of 80/92 and 91/99 for Sorek and Shoket, respectively. In addition, at both plants the final effluent is disinfected by applying hypochlorite, before it is discharged to irrigation water reservoirs.

The two WWTP sampled in this work differ in several aspects that may influence the fate of SARS-CoV-2 particles or RNA. The amount of incoming bio-solids (according to raw sewage TSS in Fig. 1) differs between the WWTP’s, which may have affected the viral load due to interactions of the virus with the biosolids. The organic loading also differs, possibly affecting viral RNA stability and the reduction of infectivity. The average removal rates of TSS (73), COD (51) and BOD (47) measured in Sorek’s primary settler were higher than those measured in Shoket’s (34, 18, 8 respectively), could affect the removal rate and detection limit of the virus at this step. Moreover, the steps in the WWT process differ between the two plants, as tertiary treatment comprising low-rate sand filtration operates only in the Shoket WWTP (Figure 1B). This step reduced TSS by ca. 50% in Shoket and could have contributed to the removal of virus particles / viral RNA attached to suspended material. Finally, operation of the chlorination step also differs between plants. In Sorek (Fig. 1A), a constant dose of hypochlorite was added (1.5 mg·L^-1^ as Cl_2_), while in Shoket the dose was adjusted to maintain the *residual* chlorine level required by the Israeli regulations (1 mg·L^-1^ as Cl_2_). Indeed, the similarities and differences the plants are reflected in the concentration of SARS-CoV-2 copies that we measured in the different treatment steps.

SARS-CoV-2 RNA (translated to CoV-2 copies per ml; see calibration curve in SI, section C2) was detected in all WW treatment stages (Figure 2). This finding has important implications regarding the safety of plant operators, since human infectivity through the WW route, e.g. by liquid aerosols, has not as yet been ruled out.^6,29^ To increase the sensitivity of the analysis, we concentrated the water samples using ultrafiltration with sterile dialysis membranes, thus lowering the effective detection limit (2000 copies·mL^-1^ for non-concentrated samples) by a factor of 17-96. This pre-concentration technique, however, was not applied to the raw sewage due to the high concentration of bio-solids. Therefore, we were able to detect SARS-CoV-2 RNA after the primary settler for all samples, while the concentration of CoV-2 copies in the raw sewage entering Sorek WWTP (Figure 2A) during April was below the detection limit (2000 copies·mL^-1^). This increased detection sensitivity has implications towards monitoring viral RNA in WW for setting up an early warning system. In Shoket WWTP, SARS-CoV-2 RNA was detected in all samples both in the raw sewage and after primary settling (Figure 2B). Unexpectedly, the number of CoV-2 copies sampled on April 27^th^ was higher than that sampled on July 14^th^ for the raw sewage, but lower for the primary effluent. This discrepancy is likely due to the variability in both the amount and type of bio-solids in the raw sewage, which can in turn affect the extent of SARS-CoV-2 attachment to the bio-solids and removal through sedimentation thereof.^7,22,42^ Thereby, when setting up a WBE study targeting SARS-CoV-2, one should consider sampling both the raw sewage and the primary effluent, which can be pre-concentrated to increase the sensitivity.

**Figure 2.**
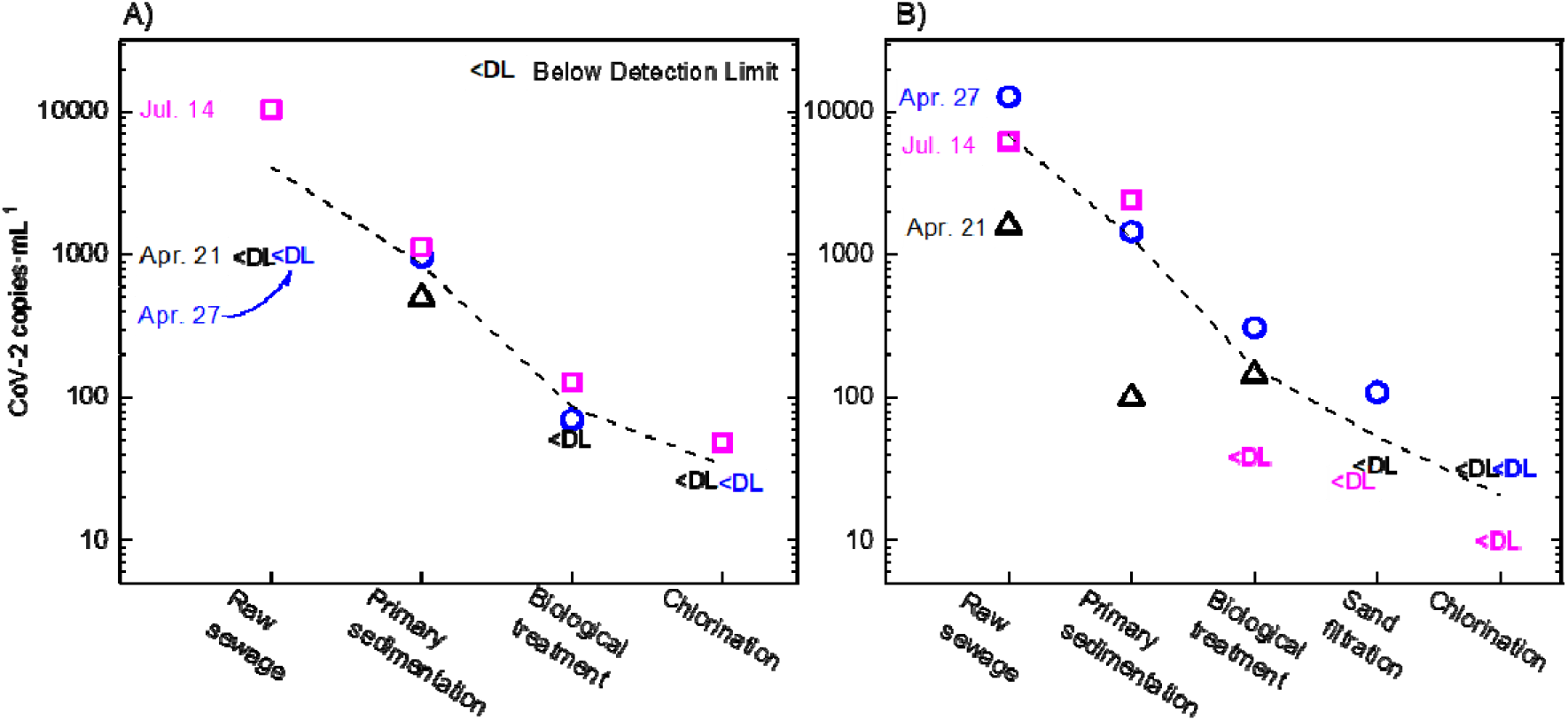
Number of CoV2-copies (N1 gene identified by RT-PCR) in the different process stages in (A) Sorek; and (B) Shoket wastewater treatment plants, at three different dates. Datapoints below detection limit (<DL) are shown at the average between 1 copy to the number of copies marking the detection limit. The dashed line is an average between 3 dates (including the <DL datapoints) to guide the eye.

The ultrafiltration pre-concentration approach (see methods and SI) enabled us to detect low concentrations of CoV-2 copies along the treatment process and assess the virus removal rate in the different WWTP stages (Figure 2). Approximately 1 Log removal was observed after primary sedimentation in all cases, indicating that most of the shed CoV-2 RNA was attached to large bio-solids. This is similar to observations^7,22,42^ for other viruses in USA, where, the settled sludge (instead of the treated effluent) was reported to contain most of the viral loads. Organic matter degradation in the biological treatment step and the subsequent secondary settling provided further ∼1 Log removal in most cases. In most WWTP’s worldwide, the second settler is the final treatment step, following which the secondary WW effluent is discharged to the environment. Our results indicate that this process can achieve an overall ∼2-Log removal of CoV-2 RNA, however the virus could still be detected in the secondary effluent.

Therefore, tertiary treatment (e.g. sand filtration) and disinfection may be needed for ensuring virus-free effluent. In Shoket WWTP (Figure 2B), sand filtration further reduced CoV-2 concentrations, once from ∼100 copies·mL^-1^ to below detection limit (April 21^st^); and once from ∼300 to ∼100 copies·mL^-1^ (Figure 1B). Virus removal in this case could be facilitated by adsorption to retained (larger) suspended particles (>10 µm) or by adsorption to the sand particles as illustrated in Fig. 1. It is noted that flocculation prior to filtration – not currently applied in Shoket – could potentially achieve higher TSS and virus removal. The subsequent addition of hypochlorite can inactivate coronaviruses ^43^ and significantly reduce the amount of SARS-CoV-2 RNA.^44^ Indeed, in Shoket WWTP (Figure 2B), the chlorination step reduced the CoV-2 copies below the detection limit, i.e. < 40 copies·mL^-1^ in the two samples collected on April and <10 copies·mL^-1^ in the one collected on July (lower detection limit stems from a higher sample pre-concentration factor). A reduction of the virus to below detection limit after chlorination was also recorded in Sorek WWTP for samples collected in April (Figure 2A). However, in July low concentration of CoV-2 RNA was detected even after chlorine application. It is possible that SARS-CoV-2 was detected after chlorination since the Shoket WWTP control hypochlorite dosage to achieve a target residual chlorine of 0.5-1 mg·mL^-1^, while the Sorek WWTP simply dose 1.5 mg·mL^-1^ hypochlorite with no consideration of residual chlorine. This approach can lead to incomplete oxidation, especially when COD is high compared to the amount of hypochlorite dosed, as in this case. These results indicate that although chlorination is effective in removing the virus, proper operation of this step, i.e. sufficient dosing to meet chlorine demand are required to ensure complete removal of SARS-CoV-2 traces.

## IMPLICATIONS AND LIMITATIONS

The results presented in this work have several significant consequences towards WW monitoring, and operating for the prevention of COVID-19 spread through WW effluents. At two different occasions we found that sampling and concentrating the primary effluent, rather than the raw sewage, increase the sensitivity of SARS-CoV-2 RNA detection, due to pre-concentration by ultrafiltration. Therefore, sampling large volumes of the primary effluent and applying pre-concentration should be considered when setting up early warning monitoring systems in WWTPs. We also note that due to the high variability of bio-solids in sewages, public health risk assessments may benefit from analysing both the raw sewage and the primary effluent. We stress that more research is needed to support these preliminary results and that improvements in the extraction and quantification methods may also contribute to advance early warning and WBE.

The detection of SARS-CoV-2 through the primary and secondary treatment calls for reinforcement of suitable safety measures to protect WWTP operators. Exposure to the virus can occur in many ways, for example through aerosols released due to the active aeration in the biological treatment rectors. Handling samples in indoors field labs for routine wastewater testing may also promote exposure. Since aerosols were shown to likely be the major infection route,^28,45^ particle filtering masks such as N95 would minimize the risk substantially. We stress that an infectivity study for the different treatment stages, not performed in this work, is needed for supporting these conclusions. Until then however, proper protection and routine testing of workers in the entire WW sector is recommended.

Furthermore, our results indicate that secondary treatment alone, as applied in most of the world today, is not sufficient for preventing the dissemination of SARS-CoV-2 to the environment through the secondary effluent. Such dissemination can lead to increased risk of infection when the water is discharged in populated areas or reused e.g. for spray irrigation. Perhaps more importantly, uncontrolled discharge of SARS-CoV-2 to the environment may increase virus accumulation in animal reservoirs (reverse zoonosis),^46^ thus impede epidemic control efforts through secondary zoonosis.^47^ Moreover, simultaneous exposure of animals to virus particles and anti-viral drugs may trigger the development of drug-resistant mutated virus.^48^ It is therefore essential to provide tighter control of SARS-CoV-2 dissemination into the environment and to further study its impact. According to our findings, tertiary treatment adds an additional barrier, and disinfection of secondary or tertiary effluents by chlorine provides an additional effective control measure. However, to ensure the effectivity of chlorination, meeting chlorine dosage demand and monitoring residual chlorine are likely to be critical. Overall, our results highlight the need for further research on the occurrence and infectivity of SARA-CoV-2 in wastewater effluent and calls for a more cautious WW management approach in these times of pandemic.

## Data Availability

All relevant data is given in the manuscript and in the Supplementary Information file

## ACKNOWLEDGEMENTS

We would like to thank funding from Ben Gurion University, The Corona Challenge Covid-19 (https://in.bgu.ac.il/en/corona-challenge/Pages/default.aspx) and funding from the Israeli ministry of Health. We thank the personnel of the Sorek and Shoket wastewater treatment plants for their support.

## Notes

### Competing Interest Statement

The authors have declared no competing interest.

### Author Declarations

No oversight was required

