## Supplementary Information for "Tracking SARS-CoV-2 RNA through the wastewater treatment process"

### Contents

**Topic**

A. Chemicals and materials
B. Target WWTPs
C. Sample processing
   A. Sewage sampling and concentration
   B. Quantification of SARS-CoV using calibration curves
D. References

### A. Chemicals and materials

All chemicals and reagents, unless specified otherwise, were of analytical grade. Milli-Q grade deionized water (18 MΩ-cm) was used wherever needed. FX100 dialysers (Fresenius Medical Care, Bud Hamburg, Germany) with an effective surface area of 2.2 $m^2$, ultrafiltration coefficient of 73 $mL \cdot h^{-1} \cdot mmHg^{-1}$ and pore size of 3.3 nm [1,2] were used for sample concentration in this study. Phosphate Buffered Saline (PBS) was prepared using phosphate buffered saline tablets purchased from Sigma-Aldrich-Merck, Israel, Commercial 10% w/w sodium hypochlorite (NaOCl) solution, used for sterilization, and ethanol (99.99%) were purchased from Romical chemicals and laboratory equipment.

### B. Target wastewater treatment plants (WWTP)

The locations of the target WWTPs (Figure S1), the corresponding COVID-19 active cases (Figure S2) in those areas and the basic physical properties of the collected WW (Table S1) are given below.

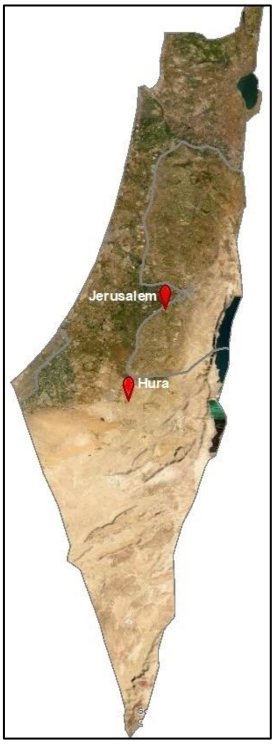

**Figure S1. WWTPs sampled in this study.** The map represents Israel. Each pin (red) corresponds to one sampling location (Shoket WWTP in Hura and Sorek WWTP in Jerusalem) Further information about the characteristics of the sampled WWTPs on the days of sampling is detailed in Table S1.

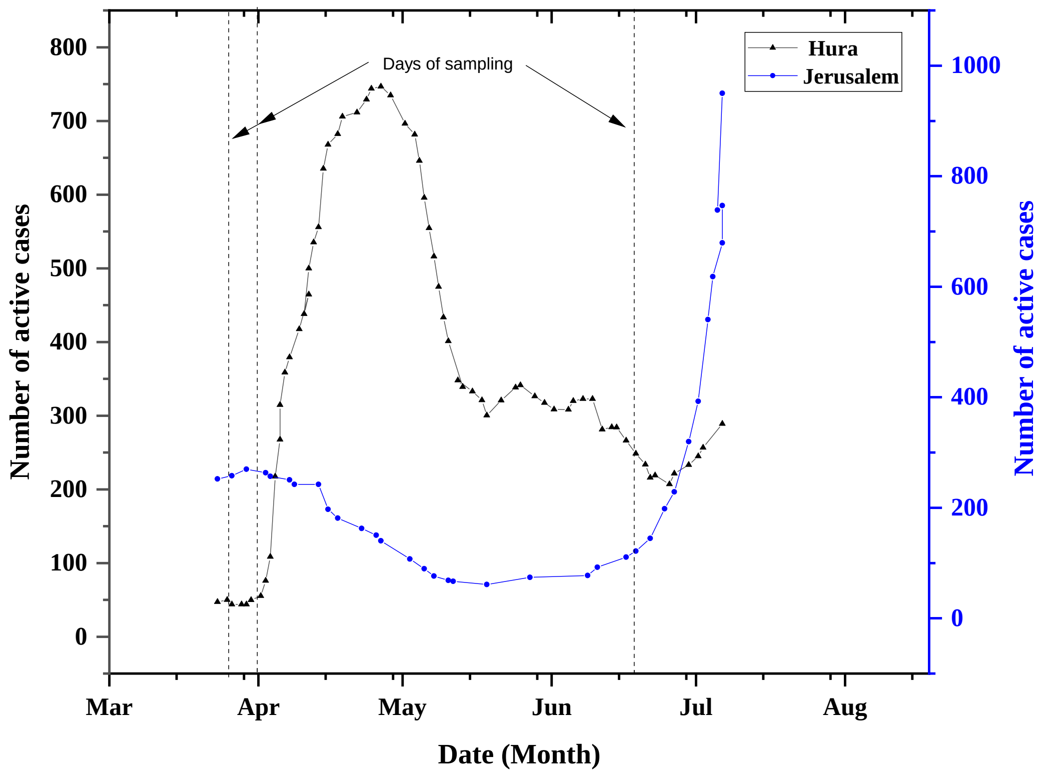

**Figure S2. Number of COVID-19 active cases in the areas served by the targeted WWTPs.**[3] Numbers of cases are per 100,000 residents. Number of active cases is calculated by removing deaths and recoveries from total cases**.**

**Table S1. Characteristics of the Targeted wastewater treatment plants (WWTPs) on the days of sampling.**

| WWTP | sampling date | average temperature (°C) | wastewater collection point | BOD ($mg \cdot L^{-1}$) | COD ($mg \cdot L^{-1}$) | TSS ($mg \cdot L^{-1}$) | residual chlorine ($mg \cdot L^{-1}$) | HRT (h)* |
|---|---|---|---|---|---|---|---|---|
| **Shoket WWTP** | 21st April, 2020 | 22°C | influent | - | - | 434 | - | |
| | | | primary sedimentation | 260 | 1194 | 290 | - | 3.5 |
| | | | secondary sedimentation | 5 | 62 | 6 | - | 8.5 |
| | | | sand filtration | 5 | 61 | 3 | - | 3.5 |
| | | | chlorine disinfection | - | - | - | 2.3 | 0.5 |
| | 27th April, 2020 | 22°C | influent | - | - | 422 | - | |
| | | | primary sedimentation | 360 | 613 | 300 | - | 3.5 |
| | | | secondary sedimentation | 4 | 46 | 2 | - | 8.5 |
| | | | sand filtration | - | - | - | - | 3.5 |
| | | | chlorine disinfection | - | - | - | 1.7 | 0.5 |
| | 14th July, 2020 | 29°C | influent | 260 | 357 | 266 | - | |
| | | | primary sedimentation | 240 | 294 | 160 | - | 3.5 |
| | | | secondary sedimentation | 1 | 43 | 8 | - | 8.5 |
| | | | sand filtration | 5 | 44 | 3 | - | 3.5 |
| | | | chlorine disinfection | - | - | - | 2.2 | 0.5 |
| **Sorek WWTP** | 21st April, 2020 | 22°C | influent | 373 | 812 | 570 | - | |
| | | | primary sedimentation | 232 | 359 | 117 | - | 1.5 |
| | | | secondary sedimentation | 10 | 76 | 11 | - | 7.9 |
| | | | sand filtration | -- | -- | -- | -- | |
| | | | chlorine disinfection | - | - | - | - | |
| | 27th April, 2020 | 22°C | influent | 485 | 885 | 562 | - | |
| | | | primary sedimentation | 329 | 519 | 221 | - | 1.5 |
| | | | secondary sedimentation | 17 | 61 | 18 | - | 7.9 |
| | | | sand filtration | -- | -- | -- | -- | |
| | | | chlorine disinfection | - | - | - | - | |
| | 14th July, 2020 | 29°C | influent | 447 | 963 | 788 | - | |
| | | | primary sedimentation | 135 | 421 | 155 | - | 1.5 |
| | | | secondary sedimentation | 18 | 113 | 122 | - | 7.9 |
| | | | sand filtration | -- | -- | -- | -- | |
| | | | chlorine disinfection | - | - | - | - | |

WWTP characteristics were retrieved from the plants' operators. '-'→ data not available; '--'→ treatment process does not exist in the plant; COD: Chemical Oxygen Demand; BOD: Biological Oxygen Demand; TSS: Total Suspended Solids; HRT: Hydraulic Retention Time (*approximate average timing).

### C. Sample processing

#### *C1. Sewage sampling and concentration*

The volumes of collected WW samples along with their corresponding concentration factors (wherever applicable) are given in Table S2. The complete method of WW sample collection to RT-PCR detection of SARS-CoV, as followed in the present work, is pictorially presented in a stepwise manner in Figure S3.

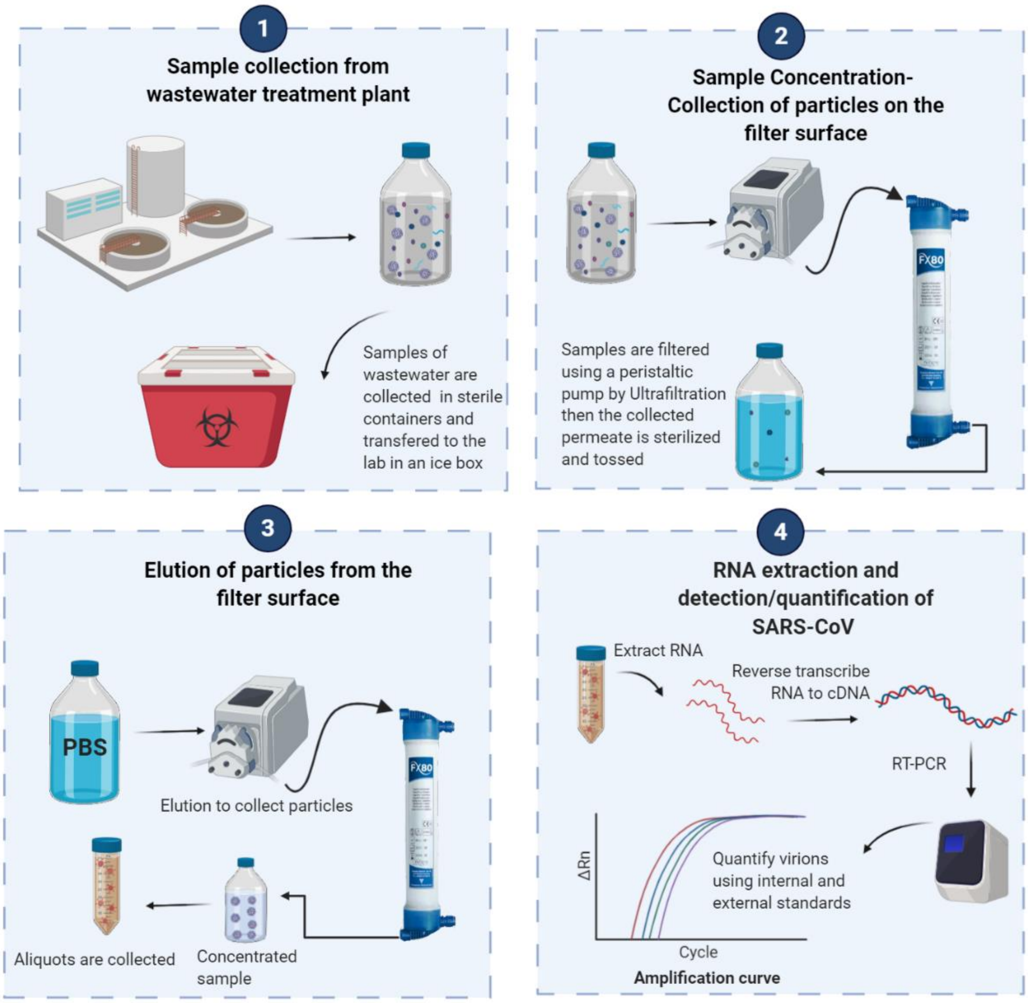

**Figure S3. Method used in quantifying SARS-CoV-2 in WW.** Scheme is created using BioRender[4]. *Step 1* – WW collections in sterilized plastic containers, S*tep 2* – concentration/volume reduction by ultrafiltration, *Step 3*- elution by PBS (phosphate buffer solution) to collect the accumulated particles on the membrane surface, *Step 4* – RNA extraction and detection of SARS-CoV by RT-PCR.

**Table S2. Concentrations of WW samples before and after concentrating by ultrafiltration method.**

| WWTP | sampling date | wastewater collection point | initial volume (L) | final volume (L) | concentration factor |
|---|---|---|---|---|---|
| Sorek WWTP | 21st April, 2020 | influent | 0.2 | 0.2 | 1 |
| | | primary sedimentation | 3.8 | 0.22 | 17.3 |
| | | secondary sedimentation | 8.5 | 0.22 | 38.6 |
| | | sand filtration | -- | - | -- |
| | | chlorine disinfection | 8.5 | 0.22 | 38.6 |
| | 27th April, 2020 | influent | 0.2 | 0.2 | 1 |
| | | primary sedimentation | 3.5 | 0.22 | 15.9 |
| | | secondary sedimentation | 8.5 | 0.22 | 38.6 |
| | | sand filtration | -- | -- | -- |
| | | chlorine disinfection | 8.5 | 0.22 | 38.6 |
| | 14th July, 2020 | influent | 0.2 | 0.2 | 1 |
| | | primary sedimentation | 9.5 | 0.27 | 35.2 |
| | | secondary sedimentation | 9.5 | 0.25 | 38 |
| | | sand filtration | -- | -- | -- |
| | | chlorine disinfection | 9.5 | 0.25 | 38 |
| Shoket WWTP | 21st April, 2020 | influent | 0.2 | 0.2 | 1 |
| | | primary sedimentation | 3.5 | 0.22 | 15.9 |
| | | secondary sedimentation | 8.5 | 0.22 | 38.6 |
| | | sand filtration | 8.5 | 0.22 | 38.6 |
| | | chlorine disinfection | 8.5 | 0.22 | 38.6 |
| | 27th April, 2020 | influent | 0.2 | 0.2 | 1 |
| | | primary sedimentation | 3.5 | 0.22 | 15.9 |
| | | secondary sedimentation | 8.5 | 0.22 | 38.6 |
| | | sand filtration | 8.5 | 0.22 | 38.6 |
| | | chlorine disinfection | 8.5 | 0.22 | 38.6 |
| | 14th July, 2020 | influent | 0.2 | 0.2 | 1 |
| | | primary sedimentation | 9.5 | 0.22 | 43.2 |
| | | secondary sedimentation | 9.5 | 0.28 | 33.9 |
| | | sand filtration | 9.5 | 0.22 | 43.2 |
| | | chlorine disinfection | 9.5 | 0.26 | 36.5 |

‘-’→ data not available; ‘--’→ treatment process does not exist in the plant.

***C2. Quantification of SARS-CoV to copy number per L***

For the calibration curve we used a plasmid contains the full SARS-CoV-2 N gene sequence as it isolated from Wuhan-Hu-1, complete genome (GenBank: NC_045512.2). We prepared serial dilutions for the plasmid and calculations for the copy number. RT-PCR amplification was performed according to user manual recommendation using CDC's N1 primers and probe set. In parallel to N1 test, each RNA sample was spiked with N gene in known concentration to rule out any inhibitors affect to the enzymatic reaction. Second quality control was done with adding MS2 phage to the lysis buffer step for RNA extraction indication (see Table s3 for RT-PCR results). To create standard curve, we performed linear regression between the log copy number and the Ct values from the RT-PCR (figure S4). Using the linear equation, we calculated copy number of N1 gene in sewage samples presented in this study.

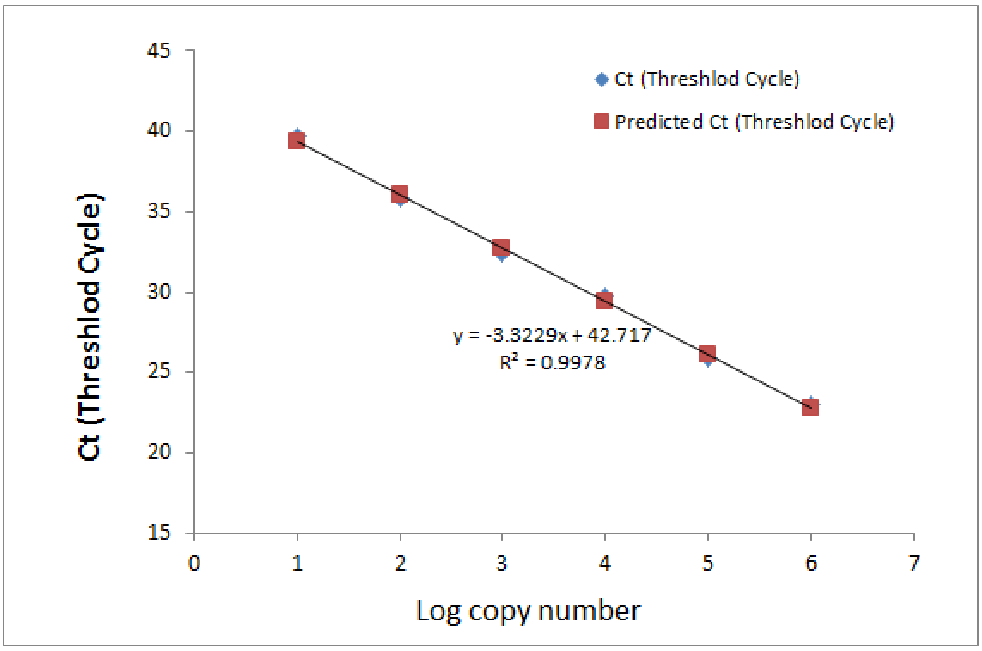

**Figure S4: calibration curve for CDC's N1 primers and probe set.** Linear regression between the log copy number of SARA-CoV-2 N gene plasmid and the threshold cycle (Ct) values from the RT-PCR amplification.

**Table S3: Ct values for detecting SARS-CoV-2 in WW samples.**

| WWTP | date | wastewater collection point | concentration factor | N1 | MS2 | Spike N gene |
|---|---|---|---|---|---|---|
| Shoket | 21st April, 2020 | influent | 1.0 | 37.34 | 29.47 | 28.86 |
| | | primary sedimentation | 15.9 | 37.35 | 29.95 | 27.98 |
| | | secondary sedimentation | 38.6 | 35.58 | 28.82 | 26.54 |
| | | sand filtration | 38.6 | - | 29.37 | 26.35 |
| | | chlorine disinfection | 38.6 | - | 29.47 | 27.04 |
| | 27th April, 2020 | influent | 1.0 | 34.41 | 30.24 | 26.31 |
| | | primary sedimentation | 15.9 | 33.58 | 30.61 | 26.91 |
| | | secondary sedimentation | 38.6 | 34.51 | 29.53 | 25.87 |
| | | sand filtration | 38.6 | 35.97 | 29.61 | 26.41 |
| | | chlorine disinfection | 38.6 | - | 29.31 | 26.21 |
| | 14th July, 2020 | influent | 1.0 | 35.42 | 28.66 | 30.1 |
| | | primary sedimentation | 43.2 | 31.47 | 30.36 | 26.15 |
| | | secondary sedimentation | 33.9 | - | 30.07 | 26.11 |
| | | sand filtration | 43.2 | - | 29.89 | 26.08 |
| | | chlorine disinfection | 36.5 | - | 29.68 | 26.33 |
| Sorek | 21st April, 2020 | influent | 1.0 | - | 29.54 | 25.94 |
| | | primary sedimentation | 17.3 | 34.97 | 30.33 | 26.31 |
| | | secondary sedimentation | 38.6 | - | 29.98 | 26.24 |
| | | sand filtration | -- | -- | -- | -- |
| | | chlorine disinfection | 38.6 | - | 29.33 | 26.65 |
| | 27th April, 2020 | influent | 1.0 | - | 29.96 | 26.17 |
| | | primary sedimentation | 15.9 | 34.16 | 29.74 | 26.71 |
| | | secondary sedimentation | 38.6 | 36.59 | 30.11 | 25.87 |
| | | sand filtration | -- | -- | -- | -- |
| | | chlorine disinfection | 38.6 | - | 29.9 | 26.84 |
| | 14th July, 2020 | influent | 1.0 | 34.7 | 28.87 | 29.89 |
| | | primary sedimentation | 35.2 | 32.83 | 29.45 | 25.43 |
| | | secondary sedimentation | 38.0 | 35.77 | 29.81 | 26.31 |

| | | | | |
|---|---|---|---|---|
| sand filtration | -- | -- | -- | -- |
| chlorine disinfection | 38.0 | 37.14 | 29.03 | 26.09 |

‘-’→ data not available; ‘--’→ treatment process does not exist in the plant.

(3) Taub Center. Three sectoral differences between the first and second wave of morbidity http://taubcenter.org.il/he/blog/heb-three-sectoral-differences-between-the-first-and-second-waves/.

(4) BioRender www.biorender.com.
